# Glycerol-Driven Contamination in PCR Labs: Implications for Diagnostic Accuracy and Laboratory Safety

**DOI:** 10.64898/2025.12.08.25341814

**Authors:** Md. Mahade Hasan Shetu, Farjana Akther Noor, Mazbahul Kabir, Md. Asadur Rahman, Akid Ornob, Maruf Hasan, Kaiissar Mannoor

## Abstract

**Background:** A key component of diagnostics in low- and middle-income (LMIC) nations is polymerase chain reaction (PCR). Despite its high sensitivity, PCR is vulnerable to laboratory contamination, particularly from aerosolized DNA. Glycerol, a common additive that enhances enzyme activity, may also promote aerosol generation—an issue not yet thoroughly investigated.

**Objective:** To determine whether PCR master mixes containing glycerol lead to increased aerosol-based contamination and false-positive results.

**Methods:** Using glycerol-free and 30% glycerol master mixes, SARS-CoV-2 N1,N2 and human RNaseP gene templates were pipetted 30 times in biosafety cabinets in two labs. Template-free tubes were exposed to ambient air for 1, 2, or 5 minutes and subjected to 40-cycle real-time PCR. Aerosol particle counts (<0.3 µm) were measured at each time point. Positive PCR counts and Ct values were compared between circumstances. Analysis was also done on retrospective contamination data comparing 10% and 30% glycerol-mixes.

**Findings:** Negative controls exposed to 30% glycerol containing master-mixes resulted in significantly earlier amplification (lower Ct values) and more frequent positivity compared to glycerol-free and 10% glycerol-mixes (P< 0.05 across all targets and time points). Aerosol particle counts were upto 3-fold higher in the 30% glycerol group than the 10% group (P< 0.001). Retrospective analysis confirmed significantly greater contamination in 30% glycerol incidents, with lower Ct values for all targets (P= 0.0001).

**Conclusion:** This work is the first to show that aerosol-mediated contamination is increased when glycerol is added to PCR master mixes. These findings emphasize strengthening biosafety measures and improving diagnostic accuracy in molecular laboratories, particularly in LMICs.

**Graphical Abstract:** 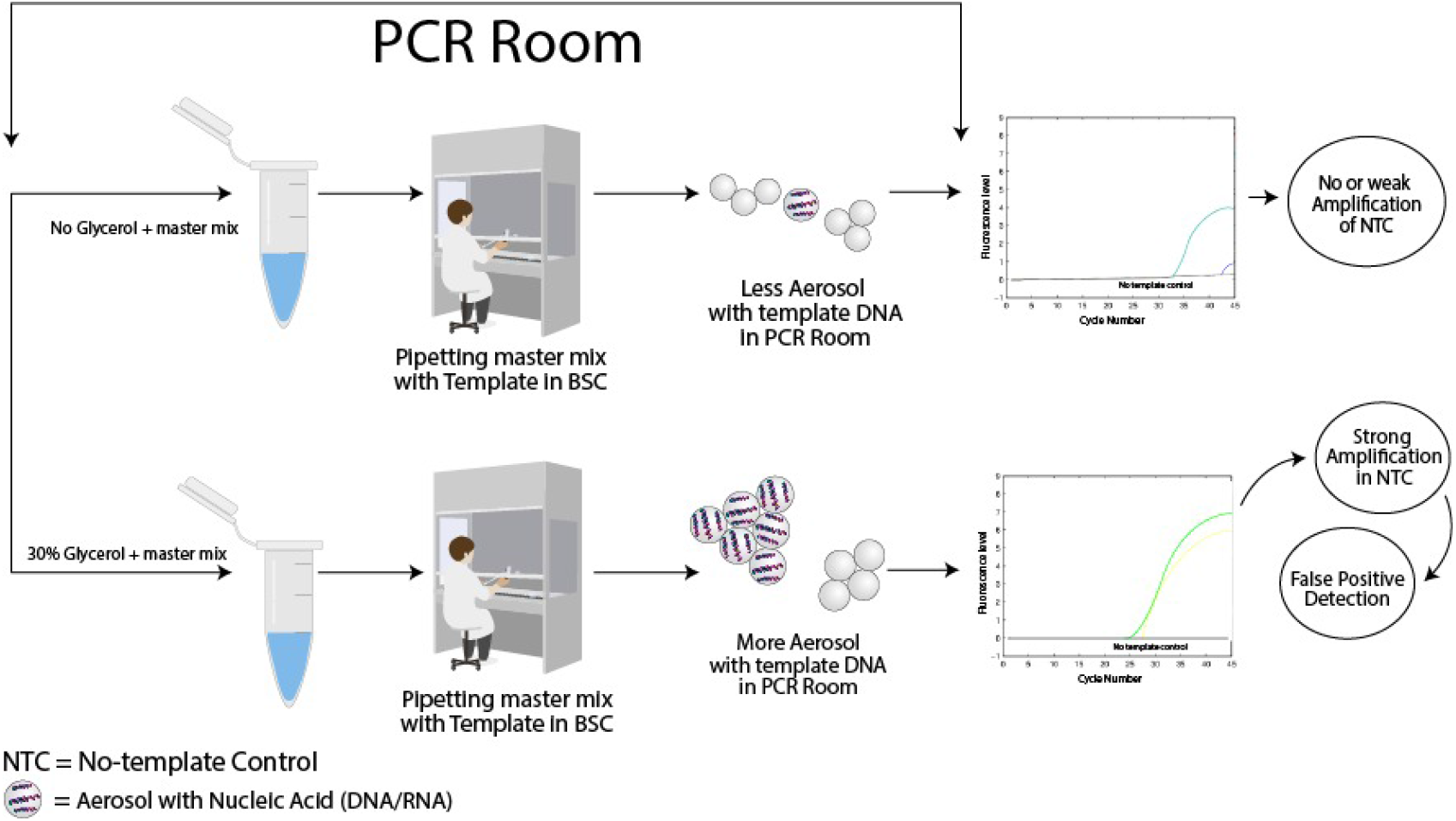

## Introduction

Laboratory medicine is fundamental to modern healthcare, serving as the backbone for disease diagnosis, therapeutic decision-making, outbreak surveillance, and public health monitoring. It plays a vital role in identifying pathogens, guiding appropriate treatment, tracking disease progression, and shaping health policies. In high-functioning systems, accurate laboratory diagnostics improve outcomes, reduce unnecessary interventions, and strengthen patient trust. However, in low- and middle-income countries (LMICs), laboratory medicine often falls short due to inadequate infrastructure, inconsistent quality control, and insufficient adherence to standardized clinical laboratory protocols.^1–4^ These systemic gaps—especially in contamination control—can result in misdiagnosis, false-positive or false-negative test results, and inappropriate therapeutic interventions. Ultimately, such diagnostic inaccuracies undermine healthcare delivery, contribute to antimicrobial resistance, and jeopardize patient safety across vulnerable populations.

Despite being highly sensitive and specific, a commonly employed nucleic acid amplification (NAA) method, polymerase chain reaction (PCR), is not flawless. In several developing nations, universal NAA standards are absent, and this is one of the causes of immense inter-laboratory variability. Environmental DNA, carryover amplicons, and positive controls are common causes of PCR laboratory contamination in LMICs. Laboratory contamination caused by aerosolized templates leads to false results due to PCR sensitivity, which compromises the test validity.^5-7^

Although the main goal of biosafety procedures is to keep pathogens in check, managing aerosolized DNA templates—whether they are plasmid-based, synthesized, or extracted—is just as crucial. The accuracy of molecular tests can be compromised by contamination with such templates, which can produce false-positive, false-negative, or inconclusive results.^5,6^

Although HEPA filters used in Class II biosafety cabinets (BSCs), such as Types A1, A2, and B1, capture 99.99% of particles ≥0.3 µm, they are less effective against tiny aerosols.^8^ False pathogen detection may also result from aerosolized DNA from positive controls or samples circulating and contaminating tubes holding negative samples or NTCs in such conditions. Therefore, in environments with limited resources, efficient aerosol management is essential to guarantee test integrity.

In this study, we report that adding glycerol to PCR master mixes may lead to an increase in aerosol generation. As demonstrated by earlier research on aerosol nuclei, glycerol is known to promote aerosol production. ^9,10^ It also plays a role in aerosol stability in electronic cigarettes.^9^ Glycerol functions as a cryoprotectant for enzymes such as Taq polymerase and increases amplification efficiency in PCR at concentrations between 15 and 50%. ^11^ Its presence, however, raises reaction viscosity, increasing the development of bubbles during pipetting or vortexing—possibly producing aerosols that could cause contamination.

We assessed SARS-CoV-2 N1, N2, and human RP gene (internal control) amplification after exposure to aerosols made from glycerol-containing versus glycerol-free master mixes in order to look into this danger. 30× pipetting of template-containing reactions was carried out in two different laboratories, and PCR tubes devoid of templates were left in room air for 1, 2, 5, or 10 minutes prior to amplification. The purpose of this investigation was to ascertain whether glycerol promotes PCR contamination and aerosol generation. This is the first study, to our knowledge, that investigates the role of glycerol in aerosol-mediated contamination in molecular diagnostics.

By identifying a hitherto unknown source of PCR contamination and offering evidence-based recommendations to increase research and diagnostic precision, lower false positives, and strengthen the dependability of molecular surveillance systems, this study has important public health implications, especially for LMICs.

## Methods

### Study design

Neither human nor animal specimens nor cell culture techniques were used in this scientific investigation. Glycerol-free (no glycerol, denoted as ‘**glycerol-**’) and glycerol-containing (30% glycerol, denoted as **‘glycerol+’**) PCR master mixes were the two independent variables in the study. A synthetic version of human RNase P (RP) was used as an internal control, and the Ct values of plasmid-based SARS-CoV-2 N1 and N2 positive controls were among the dependent variables that were examined.

We chose not to use human respiratory specimens in order to address ethical concerns. Rather, N1 and N2 targets were detected using plasmid-based positive controls that contained the SARS-CoV-2 N gene segments, and an internal control was a synthetic form of human RP. Integrated DNA Technology (IDT) provided the primers and probes that the USA CDC devised for the detection of SARS-CoV-2 N1 and N2 targets as well as human RP. These primer and probe sequences have already been described^12^. As stated in the pertinent parts, the PCR master mix utilized in this investigation was made internally, either with or without glycerol.

### Biosafety Cabinet Setup

Two molecular laboratories, one on the third floor and one on the sixth, in the same building served as the study’s locations. Class II A2 Biological Safety Cabinets (BSCs) (Model: BSC-130011A2-X, MSC Class II, Biobase, China) were installed in both facilities. Seventy percent of the filtered air was recycled back into the lab setting because there was no specialized ducting system in place. Because the pore size of the HEPA filters is 0.3 µm, aerosolized pollutants smaller than this could perhaps be recycled in the lab.

### Pre-PCR Decontamination Procedures

Prior to starting PCR studies, laboratory decontamination was carried out to reduce the possibility of contamination. The CDC (USA) decontamination guidelines for COVID-19 laboratories were followed in cleaning the work surfaces, thermal cyclers, biosafety cabinets, and other lab equipment. Non-template control (NTC) reactions were carried out using a glycerol-free PCR master mix devoid of positive templates in order to confirm the efficacy of decontamination. Prior to starting the research, the lack of target amplification in NTCs verified that the labs were free of any remaining contamination.

### Generation of Aerosolized SARS-CoV-2 and Human RP Targets

After decontamination was confirmed, controlled and repetitive pipetting inside BSCs was used to create aerosolized SARS-CoV-2 N1/N2 targets and human RP targets. Initially, glycerol-free PCR master mix without template (NTC master mix) was made in tubes and stored in the BSC of both labs with the lid closed. Aerosol was created in one lab when a glycerol-containing PCR master mix with 5000 copies/20 µL of N1/N2 plasmid and synthetic human RP DNA was pipetted 30 times inside the BSC. To produce aerosols under similar conditions, a glycerol-free master mix comprising 5000 copies/20 µL of N1/N2 plasmid and synthetic human RP DNA was pipetted 30 times inside the BSC in a different lab. Following pipetting, the NTC master mix was taken out of the BSC and left in the lab with lid opened for one, two, and five minutes before being put through a PCR reaction. In a single run, every experimental condition was replicated 32 times. Six separate runs of this type were conducted, yielding 192 repetitions for every condition. Fluorescence intensity vs. time curves were plotted for each experiment, and all PCR reactions were run for 40 cycles. The software configuration was used to examine the Ct values of positive PCR results.

## Results

### Effect of Glycerol on Laboratory Contamination: Time-Kinetics of Ct Values

To examine the effects of glycerol on lab contamination with template DNA, template-free tubes were exposed to lab air after 30× pipetting of N1, N2, and RP template-containing master mixes and tested at different time points (1, 2, and 5 minutes respectively).

Figure 1 illustrates the time-dependent kinetics of Ct values, demonstrating how repeated pipetting of glycerol-containing PCR master mix leads to progressive aerosol-mediated contamination and earlier amplification signals compared to glycerol-free formulations. Pipetting a glycerol-containing PCR master mix (30% glycerol+) inside a certified Class II A2 biosafety cabinet resulted in greater aerosol-mediated contamination compared to glycerol-free (glycerol−) mix. It is important to note that not all exposed PCR tubes yielded amplification. The Ct values presented represent only those tubes that tested positive. The proportion of positive reactions increased with exposure time, indicating a time-dependent accumulation of aerosolized DNA templates (Fig 1).

**Fig 1.**
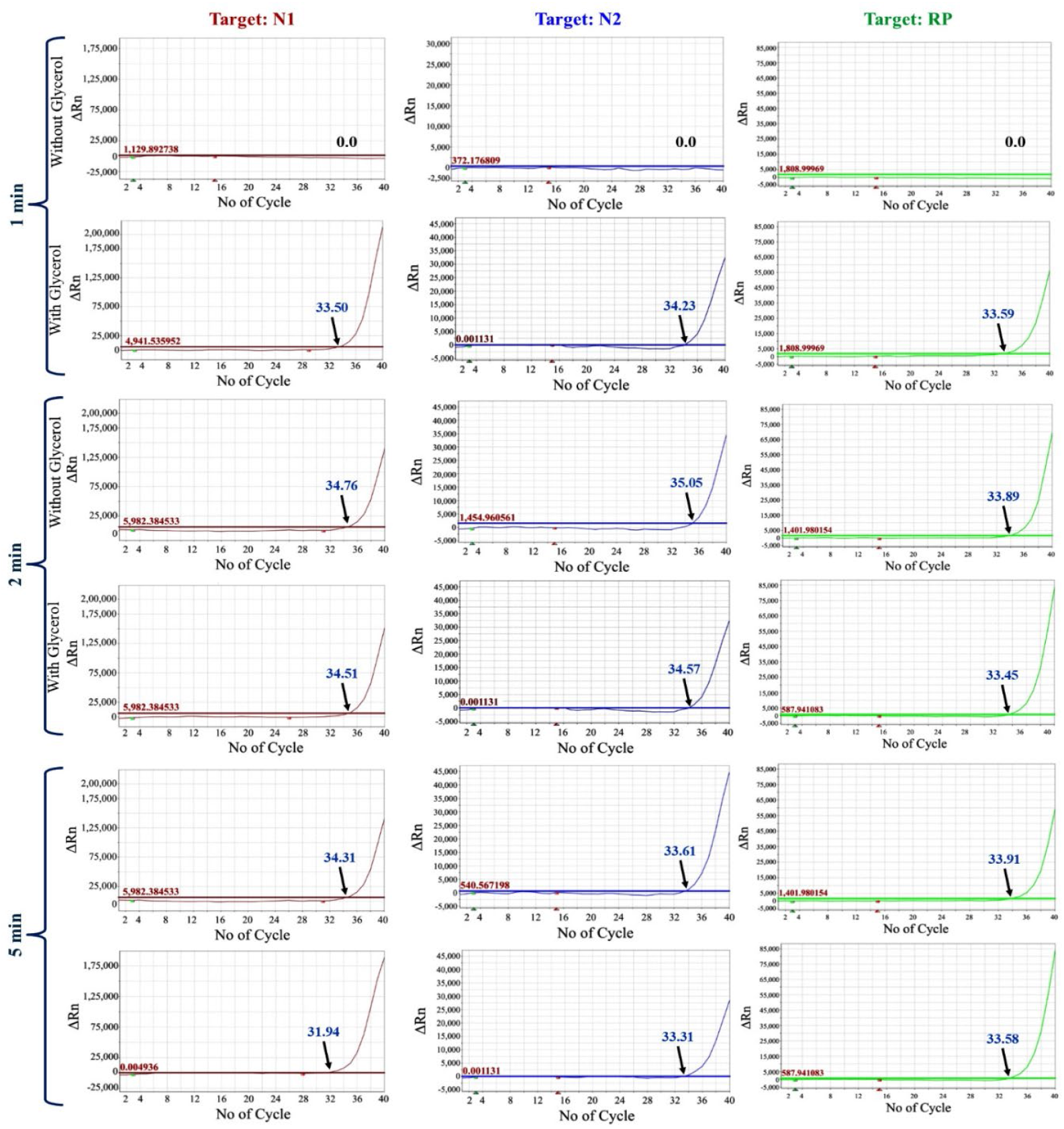
Comparison of amplification plots for SARS-CoV-2 N1, N2, and human RP (internal control) targets using PCR master mixes with (30%) and without glycerol. **Notes:** The left, middle, and right panels represent N1, N2 and RP targets, respectively. Amplification was assessed after exposing template-free master mixes to laboratory air for 1, 2, and 5 minutes, following 30× pipetting of target- containing mixes inside a biosafety cabinet.

Also, a gradual increase was found in degree of aerosol formation and subsequent contamination in terms of Ct values with longer duration of exposure. Table 1 summarizes the mean Ct values obtained from PCR reactions after repeated pipetting of DNA templates using either glycerol-free or 30% glycerol-containing master mix, as detailed in Figure 1. Following 1-minute exposure to room air, no amplification was observed in NTCs exposed in the glycerol-free setup, while all three targets amplified for the PCR positive reactions in the glycerol+ set up with mean Ct values: N1: 33.50±0.021, N2: 34.23± 0.06, RP: 33.59±0.040.

**Table 1:** Comparison of Ct values of N1, N2, and RP targets between glycerol− and 30% glycerol+ master mixes after room air exposure for 1, 2, and 5 minutes.

| Targets | 1 min<br>(mean Ct ± SD) | | | 2 min<br>(mean Ct ± SD) | | | 5 min<br>(mean Ct ± SD) | | |
| --- | --- | --- | --- | --- | --- | --- | --- | --- | --- |
|  | Glycerol<br>(-) | Glycerol<br>(+) | *P value* | Glycerol<br>(-) | Glycerol<br>(+) | *P value* | Glycerol<br>(-) | Glycerol<br>(+) | *P value* |
| N1 | 00 | 33.50<br>$\pm 0.021$ | <0.001 | 34.76<br>$\pm 0.075$ | 34.51<br>$\pm 0.098$ | 0.042 | 31.51<br>$\pm 0.0472$ | 31.21<br>$\pm 0.040$ | 0.005 |
| N2 | 00 | 34.23<br>$\pm 0.064$ | <0.001 | 35.05<br>$\pm 0.045$ | 34.57<br>$\pm 0.055$ | 0.011 | 33.61<br>$\pm 0.030$ | 33.31<br>$\pm 0.043$ | 0.001 |
| RP | 00 | 33.59<br>$\pm 0.040$ | <0.001 | 33.89<br>$\pm 0.03$ | 33.45<br>$\pm 0.035$ | 0.002 | 33.91<br>$\pm 0.051$ | 33.58<br>$\pm 0.05$ | 0.033 |

**Note:** Results represent mean Ct values ± SD from three independent experiments. In each case, 5000 copies/20 $\mu$ l of target DNA were pipetted 30 times into tubes inside a BSC to simulate aerosol formation. Template-free tubes containing corresponding master mixes were then exposed to the room and run through 40-cycle PCR. An unpaired t-test was used for statistical analysis. A p-value <0.05 was considered statistically significant.

Longer exposure led to increased contamination, evidenced by progressively lower Ct values across all three targets (N1, N2, and RP). After 2 minutes, significant reductions in Ct values were observed in samples exposed in glycerol+ set up compared to that of glycerol-free lab; the Ct values for glycerol− vs. glycerol+ were: N1=34.76 ± 0.075 vs. 34.51±0.098 (*p* = 0.0420), N2=35.05 ± 0.045 vs. 34.57±0.055 (*p* = 0.011) and RP=33.89 ± 0.03 vs. 33.45 ± 0.035 (*p =* 0.035).

This trend continued at 5 minutes, with further significant Ct decreases observed for all three targets: N1 (31.51 ± 0.047 vs. 31.21 ± 0.040, P = 0.0055), N2 (33.61 ± 0.030 vs. 33.31 ± 0.043, P = 0.0016), and RP (33.91 ± 0.051 vs. 33.58 ± 0.050, P = 0.0139). These results confirm that glycerol+ master mix increases template aerosolization, leading to earlier amplification in NTCs. Two simultaneous experiments in separate labs yielded consistent findings, reinforcing that the presence of glycerol contributes to airborne PCR contamination, with higher levels of circulating target DNA over time.

### Effect of Glycerol Concentration on Aerosol Particle Counts (≤0.3µm)

To evaluate the impact of glycerol concentration on aerosol generation, we compared particle counts (≤0.3 µm) following repeated pipetting of master mixes (MM) containing either 10% or 30% glycerol at different time points-0, 1, 2, 5 and 10 minutes. Across all time points assessed, the 30% glycerol-master mix produced significantly higher particle counts than the 10% mix (Table 2). Specifically, at 1 minute, particle counts were 48,533 ± 650.6 vs. 16,685 ± 654.0 (P = 0.0006); at 2 minutes, 25,666 ± 757.2 vs. 12,878 ± 481.3 (P = 0.0026); at 5 minutes, 19,550 ± 312.2 vs. 10,413 ± 491.0 (P = 0.0002); and at 10 minutes, 12,488 ± 457.0 vs. 6,527 ± 664.5 (P = 0.0002). These results indicate that increased glycerol content is associated with significantly greater aerosol generation during pipetting for PCR setups.

**Table 2.** Average particle counts (≤0.3 µm) in laboratory air after repeated pipetting of PCR master mixes with 10% or 30% glycerol.

| Time | Particle count ($<0.3\mu\text{m}$) for 10% glycerol-MM (particles/$\text{m}^3$) | Particle count ($<0.3\mu\text{m}$) for 30% glycerol-MM (particles/$\text{m}^3$) | *P value* |
| --- | --- | --- | --- |
| 00 min | 73800 ± 360.55 | 75900 ± 1001.66 | 0.07 |
| 01 min | 16685 ± 654.034 | 48533 ± 650.640 | 0.0006 |
| 02 min | 12878 ± 481.3089 | 25666 ± 757.187 | 0.002 |
| 05 min | 10413 ± 491.027 | 19550 ± 312.249 | 0.0002 |
| 10 min | 6527 ± 664.546 | 12488 ± 457.018 | 0.0002 |

**Notes:** Measurements were taken at 0, 1, 2, 5, and 10 minutes. Each condition was tested in separate rooms under identical settings. Values are mean ± SD from three experiments; $P < 0.05$ was considered significant.

### Effect of Glycerol on Number of Positive PCR Reactions among No-template Samples

To assess the degree of contamination severity, we also compared the frequency of positive PCR signals for the SARS-CoV-2 N1 and N2 gene targets and the human RNase P (RP) gene in no-template control (NTC) samples between master mixes with and without glycerol (glycerol+ and glycerol−, respectively). Experiments were conducted under controlled laboratory conditions, ensuring no baseline contamination. At first, 5000 copies/20uL of each target were pipetted 30 times into separate tubes containing master mixes either with or without glycerol across two independent laboratories. Subsequently, 30 template-free PCR tubes containing either glycerol+ or glycerol− master mix were exposed to laboratory air for 1, 2, or 5 minutes and subjected to 40-cycle real-time PCR.

The presence of glycerol in the master mix significantly increased airborne contamination as measured by the number of false-positive PCR reactions across all time points and target genes (Table 3). At 1-minute exposure, no false-positive signals were observed in the glycerol− group, whereas the glycerol+ group showed a mean of 4 ± 0.58, 2 ± 0.54, and 2 ± 0.55 positive reactions for N1, N2, and RP, respectively (p < 0.001 for all comparisons). Similar trends were observed at 2-minute and 5-minute exposures, with statistically significant increases in the number of positive reactions in the glycerol+ condition compared to glycerol− (e.g., N1: 2 ± 0.51 vs. 7 ± 0.53 at 2 minutes, p = 0.012; and 6 ± 0.58 vs. 10 ± 1.00 at 5 minutes, p = 0.023).

**Table 3.**
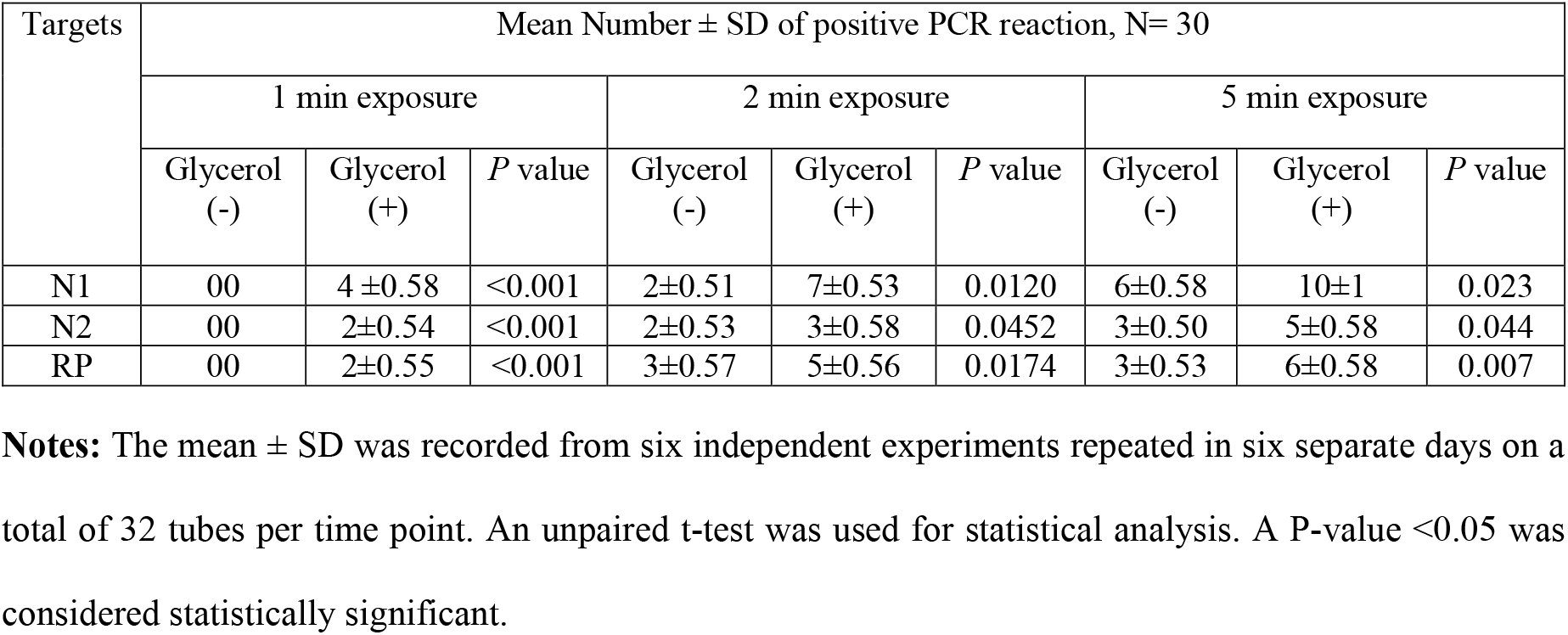
Difference in time-kinetics of the numbers of positive PCR reactions for N1, N2, RP targets between glycerol (-) MM and glycerol (+) MM.

| Targets | Mean Number ± SD of positive PCR reaction, N= 30 | | | | | | | | |
| --- | --- | --- | --- | --- | --- | --- | --- | --- | --- |
|  | 1 min exposure |  |  | 2 min exposure |  |  | 5 min exposure |  |  |
|  | Glycerol (-) | Glycerol (+) | *P* value | Glycerol (-) | Glycerol (+) | *P* value | Glycerol (-) | Glycerol (+) | *P* value |
| N1 | 00 | $4 \pm 0.58$ | $<0.001$ | $2 \pm 0.51$ | $7 \pm 0.53$ | 0.0120 | $6 \pm 0.58$ | $10 \pm 1$ | 0.023 |
| N2 | 00 | $2 \pm 0.54$ | $<0.001$ | $2 \pm 0.53$ | $3 \pm 0.58$ | 0.0452 | $3 \pm 0.50$ | $5 \pm 0.58$ | 0.044 |
| RP | 00 | $2 \pm 0.55$ | $<0.001$ | $3 \pm 0.57$ | $5 \pm 0.56$ | 0.0174 | $3 \pm 0.53$ | $6 \pm 0.58$ | 0.007 |

**Notes:** The mean ± SD was recorded from six independent experiments repeated in six separate days on a total of 32 tubes per time point. An unpaired t-test was used for statistical analysis. A *P*-value $<0.05$ was considered statistically significant.

These findings indicate a **higher frequency of aerosol-induced contamination** with glycerol+ master mix, with a time-dependent increase in positive NTCs.

### Higher Glycerol Concentration in PCR Master Mix Increases Laboratory Contamination: Evidence from Retrospective Ct Analysis

Retrospective analysis confirmed that higher glycerol concentrations led to more severe PCR contamination. Table 4 summarizes retrospective data from two independent contamination incidents during working with positive controls containing 5000 copies/20 µL of N1, N2, and RP templates in a Class II A2 biosafety cabinet. One event involved 10% glycerol master mix, and the other involved 30% glycerol master mix. After each incident, template-free PCR tubes containing 30% glycerol mix were exposed to the contaminated room air for 5 minutes and then subjected to 40-cycle PCR.

**Table 4.** Comparison of Ct values between 10% glycerol-MM and 30% glycerol-MM exposure effect in PCR reaction.

| Targets | 10% glycerol<br>(Ct values, mean ± SD) | 30% glycerol<br>(Ct values, mean ± SD) | P-value |
| --- | --- | --- | --- |
| N1 | $32.72 \pm 0.025$ | $31.46 \pm 0.025$ | 0.0001 |
| N2 | $33.77 \pm 0.050$ | $32.68 \pm 0.070$ | 0.0001 |
| RP | $34.01 \pm 0.02$ | $33.33 \pm 0.055$ | 0.0001 |

**Notes:** MM, Master Mix. Ct, Cycle of thresholds. SD, Standard Deviation. RP, Human RNase P gene. Each result represents the mean Ct ± SD from triplicate runs to assess the relative contamination levels caused by the two glycerol concentrations. A P-value $<0.05$ was considered statistically significant.

As shown in Table 4, Ct values for the 30% glycerol group were significantly lower—N1: 31.46 ± 0.025, N2: 32.68 ± 0.070, RP: 33.33 ± 0.055—compared to the 10% glycerol group—N1: 32.72 ± 0.025, N2: 33.77 ± 0.050, RP: 34.01 ± 0.020. All differences were statistically significant (P = 0.0001 for each target), indicating that the 30% glycerol master mix generated more aerosolized DNA and contributed to higher levels of laboratory contamination.

## Discussion

A key component of global health diagnostics, polymerase chain reaction (PCR) is valued and considered as reliable diagnostic tool for its sensitivity and rapidity. Contamination issues persist even though PCR reagents are typically regarded as non-hazardous, especially in low- and middle-income countries (LMICs), where biosafety infrastructure is frequently lacking. Maintaining diagnostic accuracy requires locating all possible sources of contamination and putting preventative measures into action.

Because it improves DNA polymerase thermostability and lowers the denaturation temperature of double-stranded DNA, glycerol is a frequently used component in PCR master mixes. At doses ranging from 15% to 50%, their efficacy is well established^11^ Glycerol also helps prolong the shelf life of reagents by stabilizing enzymes such as Taq polymerase during freeze-thaw cycles.

Using an in-house PCR master mix along with primers and probes designed by the US CDC, we previously developed a highly sensitive multiplex real-time reverse transcription PCR (RT-PCR) assay for the detection of SARS-CoV-2.^12^ This assay simultaneously targets two regions of the viral nucleocapsid gene (N1 and N2) as well as the human RNase P (RP) gene, which serves as an internal control. The assay was robust, reproducible, and validated across multiple samples and runs, consistently producing strong amplification signals in positive controls while ensuring the absence of amplification in no-template controls (NTCs).^12^ However, after a certain period we observed a concerning trend during routine quality control assessments: unexpected amplification signals began appearing in NTCs. NTC reactions contained all PCR components except the template nucleic acid and are designed to flag contamination of any kind. The presence of amplification in NTCs raised suspicion of environmental or reagent-associated contamination.

Upon further investigation, we confirmed that the core PCR chemistry remained functionally intact, and contamination was not due to mishandling or procedural lapses, as the laboratory followed strict biosafety protocols. However, NTC amplification was rare with glycerol-free formulations but occurred consistently with glycerol-containing master mixes. This unexpected finding prompted a detailed study into the potential role of glycerol in promoting aerosol formation and subsequent template contamination.

Our study shows the addition of glycerol to the PCR master mix may enhance the sensitivity to airborne nucleic acid contamination, likely due to its hygroscopic properties facilitating the capture of aerosolized nucleic acids. These findings were reproducible across multiple experiments and laboratories adhering to strict PPE and biosafety protocols. Notably, NTCs remained positive for 2–3 days after handling glycerol-containing mixes, despite routine decontamination with 10% bleach, 70% ethanol, and UV exposure. The issue was not merely procedural—it suggested a fundamental, under-recognized vulnerability in PCR assay design that could have implications for diagnostic accuracy, particularly in settings where environmental control is limited.

The biosafety cabinet (BSC) setup likely contributed to this issue. In our Class II A2 BSCs, 70% of HEPA-filtered air was recirculated into the room. As HEPA filters partially trap particles smaller than 0.3 µm, DNA-containing aerosols may have re-entered the workspace, contaminating subsequent PCR setups.

Such contamination has critical implications. Inaccurate or false-positive results would undermine diagnostic system validity, squander valuable resources, and affect clinical decision-making inappropriately. These issues are exacerbated in LMICs, where diagnostic capacity is already poor.

While our investigation concerned molecular diagnostics, glycerol aerosols are harmful and may pose risks that would extend beyond PCR. Glycerol finds widespread application in cryopreservation media, bioprocessing and electrophoresis buffers. Aerosol generation can be an imminent risk to laboratory personnel upon mixing with ethidium bromide or other dangerous chemicals.^13,14^ Apart from that, research has also shown that inhalation of glycerol derivatives as aerosols from combustion of biodiesel or e-cigarettes vaping can cause pulmonary and cardiovascular responses and alter the expression of circadian rhythm function genes, underlining occupational health risk assessment.^15,16^

Despite all these threats, PCR is an essential diagnostic tool in LMICs that addresses varied public health needs such as One Health monitoring, child and maternal well-being, antimicrobial resistance (AMR), infectious disease monitoring, and oncology diagnosis. Functional challenges still exist, however, because their full potential is disrupted by expensive reagents, unstable energy availability, inadequate biosafety facilities, and inadequate training of labor.

This study provides the first empirical evidence that glycerol-containing PCR master mixes contribute to an increased risk of aerosol-mediated contamination, thereby potentially leading to false-positive results. These findings have critical implications for low- and middle-income countries (LMICs), where effective AMR surveillance, epidemic response, and public health decisions all depend on diagnostic reliability. To mitigate these risks, we propose the following measures: (1) using glycerol-free or low-glycerol master mixes, particularly in high-sensitivity molecular assays, (2) improvements in biosafety cabinet design to minimize aerosol recirculation, (3) supporting local PCR reagent manufacturing to reduce cost and dependency, and (4) expanding training in biosafety-biosecurity protocols and quality control for laboratory personnel.

Implementing these strategies through targeted and feasible interventions can substantially enhance the reliability of PCR diagnostics in LMICs, reduce contamination-associated errors, and improve preparedness for emerging public health threats. Future work should explore optimal glycerol concentrations that balance reagent stability with contamination risk.

## Data Availability

All data produced in the present work are contained in the manuscript

## Declaration

### Availability of data and materials

All relevant data are within the paper. Further information is available from the authors on request.

### Consent for publication

All the authors have read and approved the paper for publication.

